# A combined strategy to detect plasma samples reliably with high anti-SARS-CoV-2 neutralizing antibody titers in routine laboratories

**DOI:** 10.1101/2021.03.15.21253588

**Authors:** Bastian Fischer, Christoph Lichtenberg, Lisa Müller, Jörg Timm, Johannes Fischer, Cornelius Knabbe

## Abstract

The determination of anti-SARS-CoV-2 neutralizing antibodies (NAbs) is of interest in many respects. High NAb titers, for example, are the most important criterion regarding the effectiveness of convalescent plasma therapy. However, common cell culture-based NAb assays are time-consuming and feasible only in special laboratories. Our data reveal the suitability of a novel ELISA-based surrogate virus neutralization test (sVNT) to easily measure the inhibition-capability of NAbs in the plasma of COVID-19 convalescents. We propose a combined strategy to detect plasma samples with high NAb titers (≥ 1:160) reliably and to, simultaneously, reduce the risk of erroneously identifying low-titer specimens. For this approach, results of the sVNT assay are compared to and combined with those acquired from the Euroimmun anti-SARS-CoV-2 IgG assay. Both assays are appropriate for high-throughput screening in standard BSL-2 laboratories. Our measurements further show a long-lasting humoral immunity of at least 11 months after symptom onset.

## Introduction

Severe acute respiratory syndrome coronavirus 2 (SARS-CoV-2) was first identified at the end of December 2019 in Wuhan, Hubei Province, China [1]. Sequencing analysis from the lower respiratory tract revealed the new coronavirus early as a causative agent of the Coronavirus disease 2019 (COVID-19) [2]. The infectious disease became a worldwide pandemic and has claimed millions of lives so far. While most infections are mild or even asymptomatic, the estimated infection fatality rate across populations is 0.68 % (0.53 – 0.82 %) [3]. While vaccines are promising concerning the formation of an active immunization against the virus, passive immunization can be achieved by an early treatment of SARS-CoV-2-infected individuals with the plasma of COVID-19 convalescent donors [4]. The most important criterion regarding the effectiveness of the convalescent plasma (CP) therapy is a high concentration of anti-SARS-CoV-2-neutralizing antibodies (NAbs) [5]. However, the determination of NAbs is time-consuming and can, due to the use of live authentic SARS-CoV-2 viruses, only be performed in high safety Biosafety Level 3 (BSL3) cell culture laboratories [6]. In order to select the appropriate CP, therefore, the concentration of total anti-SARS-CoV-2-binding antibodies (BAbs) is often considered, for which different serological assays are commercially available. Previous studies have revealed a moderate correlation between anti-spike IgG levels and NAb titers determined in a cell culture-based assay [7]. However, no statement about the antibody functionality can be made by the determination of general BAbs. Therefore, the usage of functional NAb assays is indispensable to assess the protective humoral immunity against SARS-CoV-2 after natural infection or vaccination.

We compared the results of a novel enzyme-linked immunosorbent assay (ELISA)- based surrogate virus neutralization test (sVNT) for the detection of anti-SARS-CoV-2 NAbs with those of a cell culture assay. The results were additionally correlated with total anti-SARS-CoV-2 IgG BAb ratios determined using the serological Euroimmun test. Based on our findings, we suggest a combined strategy to reliably detect samples with high NAb titers, while strongly reducing the number of false-positive, low-titer samples.

## Results

### Comparison of the sVNT and a cell culture assay for the determination of anti-SARS-CoV-2 NAbs

A total of 108 residual blood samples of 98 COVID-19 convalescents, donated in the period between April 2020 and January 2021, were tested for the presence of anti-SARS-CoV-2 NAbs using two different assays. All donors underwent a medical examination before donation. Samples were collected in accordance with the German Act on Medical Devices for the collection of human residual material. Ethical approval was obtained from the ethical committee of the HDZ NRW in Bad Oeynhausen (Reg. No. 670/2020).

The sVNT ELISA from GenScript (Piscataway Township, USA) is designed to mimic the virus–host interaction using a purified receptor-binding domain (RBD) protein and immobilized cell surface receptor, angiotensin converting enzyme-2 (ACE2). Due to horseradish peroxidase-conjugated RBD, the absorbance of a sample can be measured at 450 nm and is inversely proportional to the NAb titer of the respective specimen. The experimental procedure was performed as specified by the manufacturer [8]. The cell culture assay was performed as previously described [9]. In brief, a virus stock solution with the SARS-CoV-2 NRW-42 isolate (EPI ISL 425126 [10]) was added to a final concentration of 1000 TCID50/well to heat-inactivated and diluted plasma samples. The plasma neutralization titer was determined by microscopic inspection as the highest plasma dilution without a virus-induced cytopathic effect. All samples were tested in duplicate. Results of both assays show a good correlation and NAbs were detected in all donors, as shown in Figure 1. The manufacturer’s specified cutoff value of 20 % was used for the ELISA-based surrogate assay.

**Figure 1:**
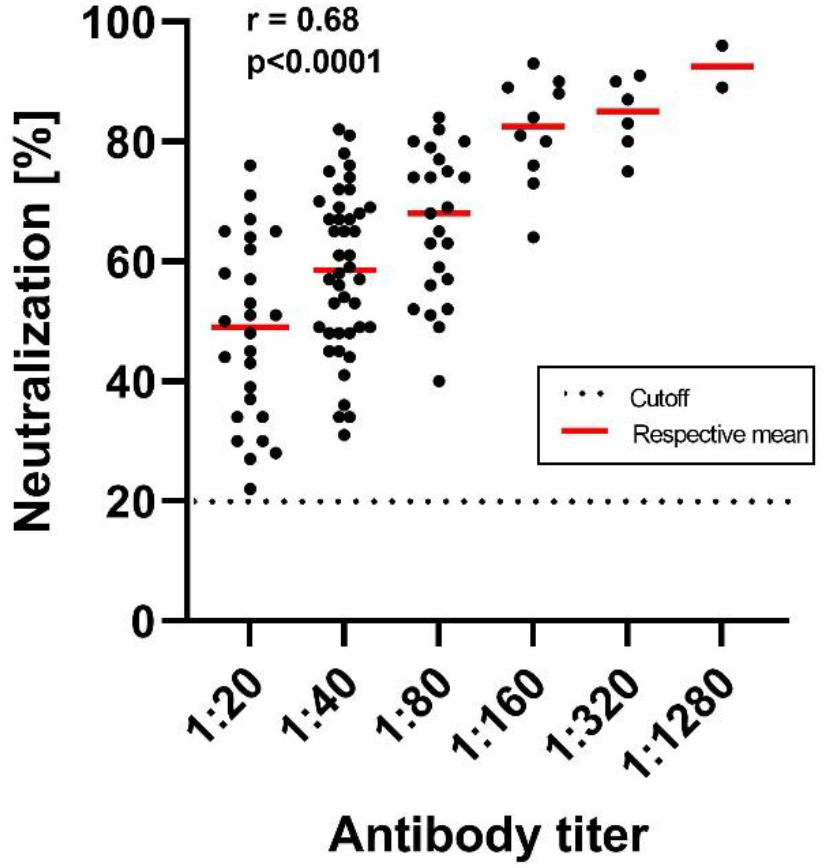
Comparison of the results obtained from the sVNT ELISA and the cell culture assay for the determination of anti-SARS-CoV-2 neutralizing antibodies. The dotted horizontal line symbolizes the positive cutoff (20 %) of the sVNT assay specified by the manufacturer. The correlation coefficient was determined using one-way ANOVA.

### Correlation of anti-SARS-CoV-2 IgG NAbs and BAbs

Residual blood samples were additionally tested for the presence of total anti-SARS-CoV-2 IgG BAbs directed against domain S1 of the viral spike protein using the serological ELISA of Euroimmun (Lübeck, Germany). Semiquantitative results were calculated as a ratio of the extinction of samples over the extinction of a calibrator. Measurements were fully automated, according to the manufacturer’s protocol, using the Euroimmun Analyzer I system. A good correlation of the values determined in the cell culture NAb and Euroimmun assay was generally observed (r = 0.71), with occasional samples revealing high NAbs despite comparatively low anti-SARS-CoV-2 IgG ratios. All convalescents tested showed SARS-CoV-2 IgG seroconversion (Figure 2).

**Figure 2:**
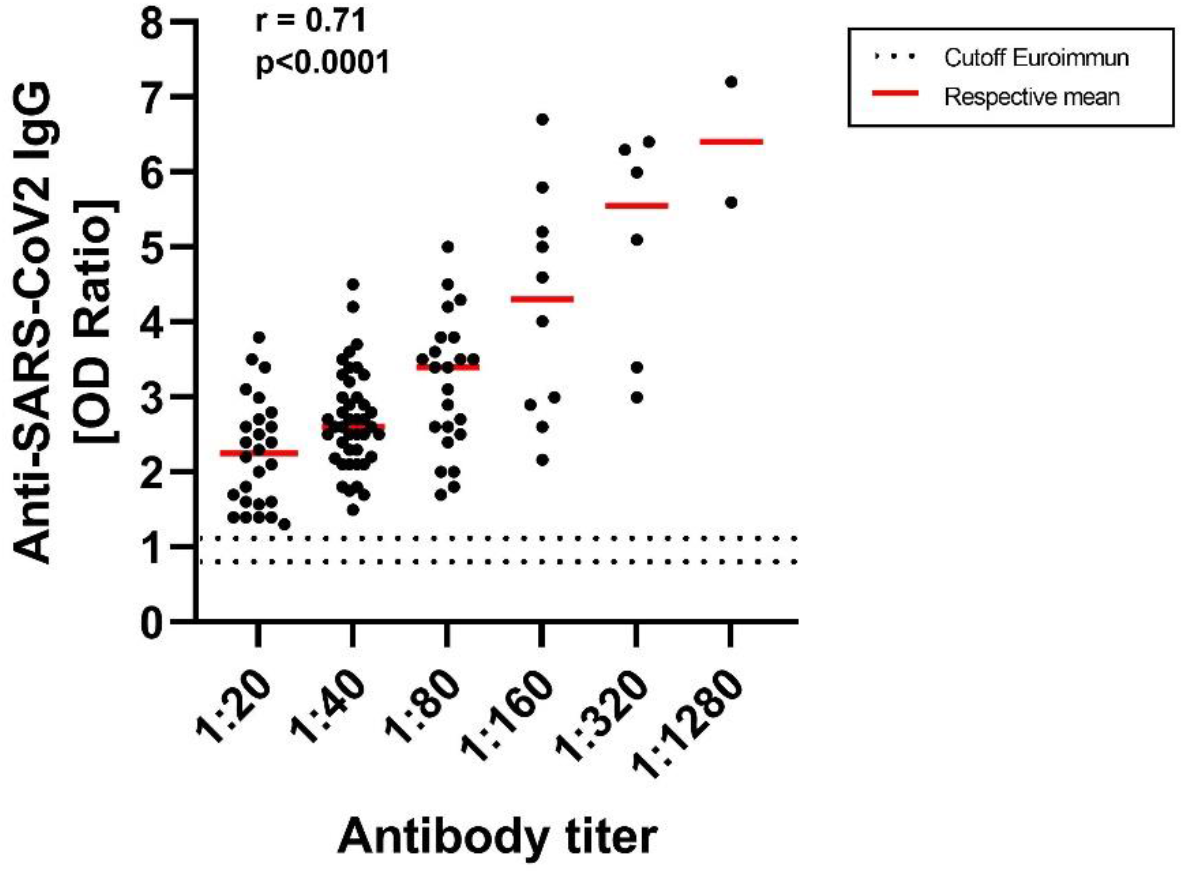
Comparison of the cell culture neutralizing antibody (NAb) assay and the semiquantitative Euroimmun assays for the detection of anti-SARS-CoV-2 IgG binding antibodies (BAbs). The dotted horizontal lines symbolize the positive (OD ratio: 1.1) and the equivocal (OD ratio: 0.8) cutoff of the anti-SARS-CoV-2 IgG Euroimmun assay specified by the manufacturer. All convalescents included showed SARS-CoV-2 seroconversion. The correlation coefficient was determined using one-way ANOVA.

The percentage neutralization values determined using the sVNT assay also showed a good correlation with anti-SARS-CoV-2 IgG ratios (r = 0.74). We additionally color-coded the different NAb titers determined in the cell culture assay for a better overview (Figure 3). When using a self-defined cutoff of 75 % for the sVNT assay, 94 % (17/18) of high-titer specimens (> 1:160) were identified. Nevertheless, samples with lower titers were also detected (titer 1:80: 7/23 (30 %), titer 1:40: 5/42 (12 %) and titer 1:20: 1/25 (4 %)) while only respecting this cutoff value. When using an additional cutoff of a ratio 4.6 for the anti-SARS-CoV-2 IgG ELISA, only 4 % (titer 1:80: 1/23) of samples with lower antibody titers were detected. However, this strategy also results in only 61 % of high-titer specimens (titer ≥ 1:160: 11/18) being captured.

**Figure 3:**
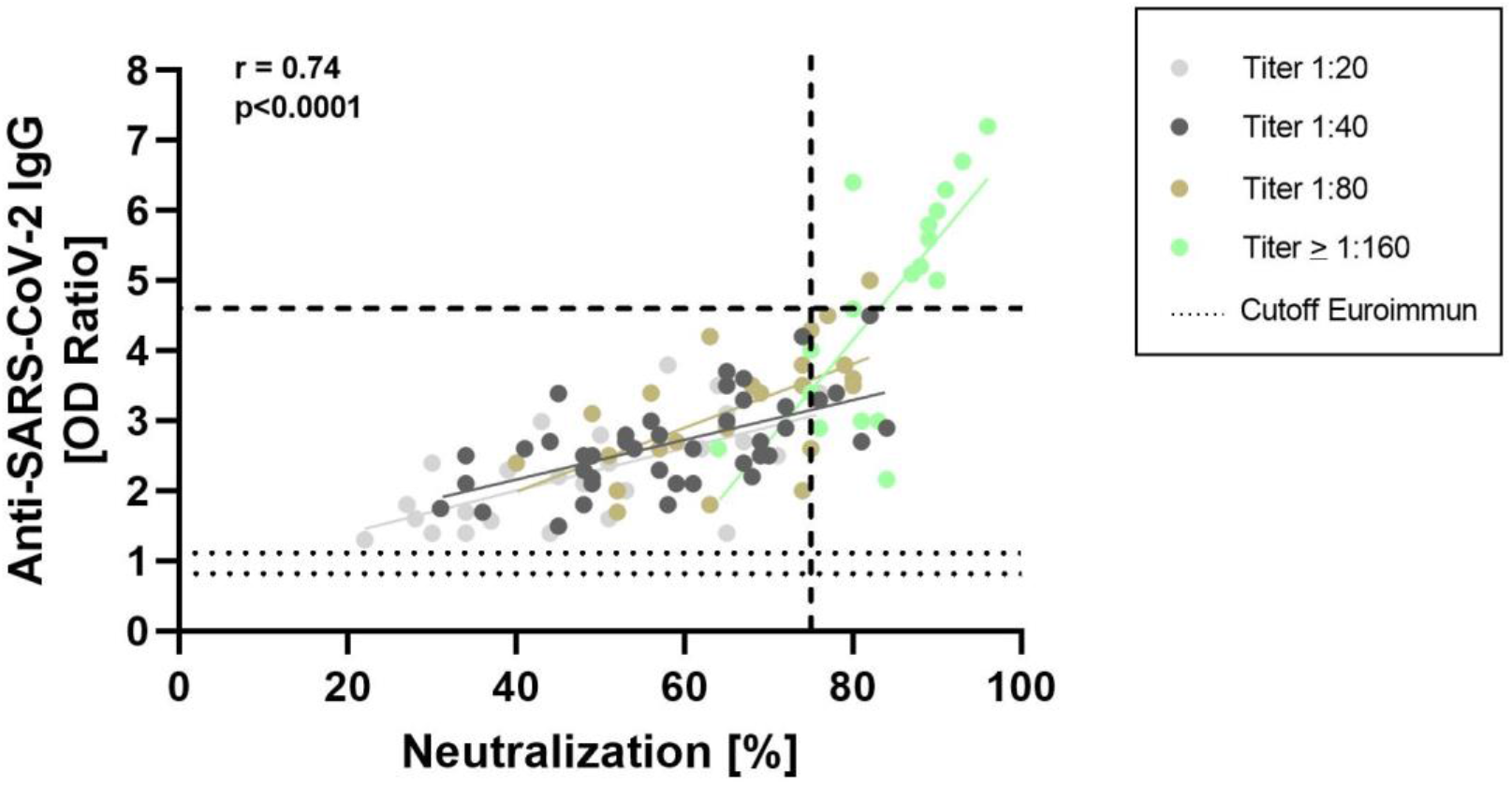
Comparison of the ELISA-based sVNT NAb assay and the serological Euroimmun assay for the detection of anti-SARS-CoV-2 IgG BAbs. The dotted horizontal lines symbolize the positive (OD ratio: 1.1) and the equivocal (OD ratio: 0.8) cutoff of the anti-SARS-CoV-2 IgG Euroimmun assay specified by the manufacturer. The dashed vertical line symbolizes our suggested positive cutoff for the sVNT assay of 75 %. Defining an additional cutoff for the Euroimmun assay at a ratio of 4.6 (dashed horizontal line) would lead to a reduced identification of samples with low antibody titers. The correlation coefficient was calculated using simple linear regression.

## Discussion

Passive immunization is a promising approach to protect SARS-CoV-2-infected individuals from a severe COVID-19 course. Data from a prospective study suggest that early treatment of infected adults with CP can prevent severe COVID-19 by up to 73 % [11]. The effectiveness of therapy depends crucially on the NAb concentration of the plasma transfused. Duan et al. showed that transfusing one dose (200 ml) of COVID-19 CP with a NAb titer of ≥ 1:160 significantly improved the clinical outcomes of ten patients suffering from COVID-19 disease [12]. The NAbs are determined standardly using cell culture-based assays, which can only be performed in special BSL3 laboratories and are very time-consuming (several days). An alternative is the novel ELISA-based sVNT neutralization assay, which has some practical advantages: It can be performed in any standard Biosafety Level 2 (BSL2) laboratory within a few hours, does not require special equipment and is feasible for high-throughput testing [8]. In addition to some standardized seroassays for the identification of total anti-SARS-CoV-2 IgG BAbs, which do not address the functionality of the antibody response, the sVNT assay has been approved by the U.S. Food and Drug Administration (FDA) as being “acceptable for use in the manufacture of high titer COVID-19 convalescent plasma.” As a qualifying criterion for therapeutic CP, the FDA recommends a ratio of ≥ 3.5 concerning the anti-SARS-CoV-2 IgG Euroimmun assay and an inhibition-value ≥ 68 % for the sVNT neutralization test. According to the FDA, NAb titers should be ≥ 1:160 when using a common cell culture assay to be adequate for therapy [13]. All 98 convalescents included in our cohort expressed anti-SARS-CoV-2 NAbs detectable in both assays, whereby results show a good correlation (Fig. 1). Assay correlation (r = 0.68) was comparable to those reported in previous studies [14, 15]. It is of note that our data also suggest long-lasting humoral immunity of at least 11 months against the new coronavirus, as the maximum period between symptom onset and donation was 323 days. This is, to the best of our knowledge, the longest reported persistence of humoral immunity against SARS-CoV-2 so far.

Results of both NAb assays showed a good correlation to those of the Euroimmun anti-SARS-CoV-2 IgG assay (cell culture assay: r = 0.71, sVNT assay: r = 0.74). By contrast, only a weak correlation between NAbs and anti-SARS-CoV-2 IgA antibodies (Euroimmun, Lübeck) was detected for both assays (cell culture assay: r = 0.55, sVNT assay: r = 0.32, see supplement). While data for the sVNT assay are lacking, a recent study of Müller et al. has already shown a good correlation between a cell culture-based NAb assay and anti-SARS-CoV-2 IgG BAb ratios determined by using the Euroimmun ELISA assay [9]. As opposed to our data, the authors also showed a good correlation to anti-SARS-CoV-2 IgA antibody results. This might be explainable by the fact that IgA-antibody levels seem to drop rapidly, whereby IgG antibodies against the virus are stably detectable for several months in individuals who have recovered from COVID-19 [16, 17]. Therefore, the correlation between NAbs and IgA antibodies depends strongly on the time post-infection.

Using the anti-SARS-CoV-2 IgG ratio as the only criterion would result in some plasmas with high NAbs not being identified, as shown in Fig. 2 and 3. Importantly, this approach also detects occasional plasmas showing relatively low NAb concentrations and would, therefore, not be appropriate for CP therapy. By using an inhibition value of ≥ 75 % as a positive cutoff for the sVNT assay, we detected 94 % (17/18) of high-titer plasmas (titer ≥1:160). However, this approach also results in the identification of a considerable number of plasmas showing lower NAb titers in the cell culture assay (Fig. 3). To avoid this, we propose a combined approach concerning qualification of the CP, which can be performed in any standard laboratory. Based on our results, using a positive cutoff of ratio ≥ 4.6 for the Euroimmun IgG assay and an inhibition value ≥ 75 % for the sVNT assay reduces the identification of “false-positive” low-titer plasmas strongly, while detecting 61 % of high-titer specimens (titer ≥ 1:160; 11/18).

In summary, based on our results, we propose a combined strategy to detect plasma samples showing high NAb titers reliably and additionally reduce the risk of identifying false-positive, low-titer specimens. Our data further reveal a long-lasting humoral immunity against SARS-CoV-2 of at least 11 months.

## Data Availability

The datasets generated during the current study are available from the corresponding author on reasonable request.

## Abbreviations

SARS-CoV-2: Severe acute respiratory syndrome coronavirus type 2
COVID-19: Coronavirus Disease 2019
NAbs: Neutralizing antibodies
Babs: Binding antibodies

